# How does ChatGPT4 preform on Non-English National Medical Licensing Examination? An Evaluation in Chinese Language

**DOI:** 10.1101/2023.05.03.23289443

**Authors:** Changchang Fang, Jitao Ling, Jing Zhou, Yue Wang, Xiaolin Liu, Yuan Jiang, Yifan Wu, Yixuan Chen, Zhichen Zhu, Jianyong Ma, Ziwei Yan, Peng Yu, Xiao Liu

## Abstract

**Background:** ChatGPT, an artificial intelligence (AI) system powered by large-scale language models, has garnered significant interest in the healthcare. Its performance dependent on the quality and amount of training data available for specific language. This study aims to assess the of ChatGPT’s ability in medical education and clinical decision-making within the Chinese context.

**Methods:** We utilized a dataset from the Chinese National Medical Licensing Examination (NMLE) to assess ChatGPT-4’s proficiency in medical knowledge within the Chinese language. Performance indicators, including score, accuracy, and concordance (confirmation of answers through explanation), were employed to evaluate ChatGPT’s effectiveness in both original and encoded medical questions. Additionally, we translated the original Chinese questions into English to explore potential avenues for improvement.

**Results:** ChatGPT scored 442/600 for original questions in Chinese, surpassing the passing threshold of 360/600. However, ChatGPT demonstrated reduced accuracy in addressing open-ended questions, with an overall accuracy rate of 47.7%. Despite this, ChatGPT displayed commendable consistency, achieving a 75% concordance rate across all case analysis questions. Moreover, translating Chinese case analysis questions into English yielded only marginal improvements in ChatGPT’s performance (P =0.728).

**Conclusion:** ChatGPT exhibits remarkable precision and reliability when handling the NMLE in Chinese language. Translation of NMLE questions from Chinese to English does not yield an improvement in ChatGPT’s performance.

## Introduction

AI (Artificial Intelligence) has gained significant influence in recent years, simulating human intelligence and cognitive processes to tackle complex problems^1^. Trained on specific datasets, AI systems enhance prediction accuracy and address complex challenges^2-4^, assisting doctors in rapidly searching through medical data, augmenting creativity, and facilitating error-free decision-making^5, 6^. ChatGPT is a Large Language Model that predicts word sequences based on context and generates novel sequences resembling natural human language. These novel sequences have not been previously observed by other AI systems ^7^.

ChatGPT shows promise in medical education, performing well in Certified Public Accountant (CPA) exams and generating accurate responses to complex inputs8. Applied in the United States Medical Licensing Examination and South Korean parasitology exams, ChatGPT demonstrates significant advancements, despite discrepancies with medical students’ scores^9^. However, ChatGPT’s proficiency relies on available training data quality and quantity in the languages, and most of them is in English. With over 1.3 billion speakers, the amount and quality of training data in Chinese language may not be comparable to English, necessitating further research into ChatGPT’s performance in Chinese medical information. The Chinese National Medical Licensing Examination (NMLE) is a legally mandated qualification for doctors^10^. This comprehensive, standardized assessment poses conceptually and linguistically challenging questions across medical domains, which makes it an excellent input for ChatGPT in clinical decision-making.

Give this background, this study aims to evaluate ChatGPT’s performance on the Chinese NMLE conducted within the Chinese context.

## Methods

### Artificial Intelligence

ChatGPT is an advanced language model that leverages self-attention mechanisms and extensive training data to deliver natural language responses within conversational settings. Its primary strengths encompass managing long-range dependencies and producing coherent, contextually appropriate responses. Nevertheless, it is essential to recognize that GPT-4 is a server-based language model without internet browsing or search capabilities. Consequently, all generated responses rely solely on the abstract associations between words, or “tokens,” within its neural network^7^.

### Input source

The official website does not release the 2022 NMLE test questions. However, a complete set of 600 questions, with a total value of 600 points, is available online (Supplemental S1) and considered as original questions. These questions are divided into four units, with each question worth one point.

The four units encompass the following areas: Unit 1 assesses medical knowledge, policies, regulations, and preventive medicine; Unit 2 focuses on cardiovascular, urinary, muscular, and endocrine systems; Unit 3 addresses digestive, respiratory, and other related systems; while Unit 4 evaluates knowledge of female reproductive systems, pediatric diseases, and mental and nervous systems.

All inputs provided to the GPT-4 model are valid samples that do not belong to the training dataset, as the database has not been updated since September 2021, predating the release of these questions, this was further confirmed by randomly spot checking the inputs. To facilitate research efforts, the 600 questions have been organized into distinct categories based on their question type and units.

1. Common Questions (n=340): These questions are distributed across all units, including Unit 1 (n=108), Unit 2 (n=82), Unit 3 (n=79), and Unit 4 (n=71). They aim to evaluate basic science knowledge in physiology, biochemistry, pathology, and medical humanities. Each question has four choices, and the AI must select the single correct answer. An example from Unit 1 is: “What type of hypoxia is likely to be caused by long-term consumption of pickled foods? A. Hypoxia of blood type B. Hypoxia of tissue type C. Circulatory hypoxia D. Anoxic hypoxia E. Hypoxia of hypotonic type.”

2. Case Analysis Questions (n=260): These questions are also distributed across all units, including Unit 1 (n=42), Unit 2 (n=68), Unit 3 (n=71), and Unit 4 (n=79). Employed in clinical medicine, it examines and evaluates patient cases through a thorough review of medical history, symptoms and diagnostic findings to determine a diagnosis and treatment plan. Each question has four choices, and the AI must select the single correct answer. An example from Unit 1 is: “A 28-year-old male complains of muscle and joint pain in his limbs three days after diving. He experienced respiratory equipment failure during diving three days ago and immediately ascended rapidly to the surface. Subsequently, he experienced symptoms such as dizziness, orientation disorder, nausea, and vomiting. After rest and oxygen inhalation, the symptoms improved, but he continued to experience persistent muscle spasms, convulsions, and joint pain in his limbs. Therefore, what is the most likely cause of the patient’s pain? A. Chronic inflammation and cell infiltration B. Stress ulcers C. Local tissue coagulative necrosis D. Increased carbon dioxide concentration in the blood E. Gas embolism in the blood vessel lumen.”

### Scoring

We assembled a dataset of NMLE questions and their corresponding answers, maintaining validity by cross-verification with senior medical professionals. This dataset was used to evaluate ChatGPT’s performance on the exam by comparing its responses to the standard answers and calculating the scores it achieved. A high score would indicate that ChatGPT effectively tackled this task.

### Encoding

To better reflect the actual clinical situation, we modified the case analysis questions to be open-ended. Questions were formatted by deleting all the choices and adding a variable lead-in imperative or interrogative phrase, requiring ChatGPT to provide a rationale for the answer choice. Examples include: “What could be the most plausible explanation for the patient’s nocturnal symptoms? Justify your answer for each option,” and “Which mechanism is most likely responsible for the most fitting pharmacotherapy for this patient? Provide an explanation for its correctness.”

However, a unique subset of questions could not be encoded in the same manner. These questions required selecting one provided choice, so we transformed them into a special form (n=3). For example, the original question, “Which can inhibit insulin secretion? A. Increased free fatty acids in blood B. Increased gastric inhibitory peptide secretion C. Sympathetic nerve excitation D. Growth hormone secretion increases” was encoded as “Can an increase in free fatty acids in the blood, an increase in gastric inhibitory peptide secretion, an increase in sympathetic nerve excitation, or an increase in growth hormone secretion inhibit insulin secretion?” This encoding was present only in Unit 1. To minimize memory retention bias, a new chat session was initiated for each inquiry.

### Adjudication

In our study, AI outputs from the two types of encoders were independently scored for Accuracy and Concordance by two physicians blinded to each other’s assessments. Scoring was based on predefined criteria (**Supplemental S2**). To train the physician adjudicators, who were not blinded to each other, a subset of 20 questions was used. ChatGPT’s responses were classified into three categories: accurate, inaccurate, and indeterminate. Accurate responses indicated that ChatGPT provided the correct answer, while inaccurate responses encompassed no answer, incorrect answers, or multiple answers with incorrect options. Indeterminate responses implied that the AI output did not provide a definitive answer selection or believed there was insufficient information to do so. Concordance was defined as when ChatGPT’s explanation confirmed its provided answer, while discordant explanations contradicted the answer. To minimize within-item anchoring bias, adjudicators first evaluated accuracy for all items, followed by concordance. Two physicians were blinded to each other’s evaluations. In cases of discrepancy, a third physician adjudicator was consulted. Ultimately, 17 items (2.7% of the dataset) required the intervention of a third physician adjudicator. The interrater agreement between the physicians was assessed using the Cohen kappa (κ) statistic for the questions (**Supplemental S3**). A schematic overview of the study protocol is provided in **Fig 1**.

**Fig 1.**
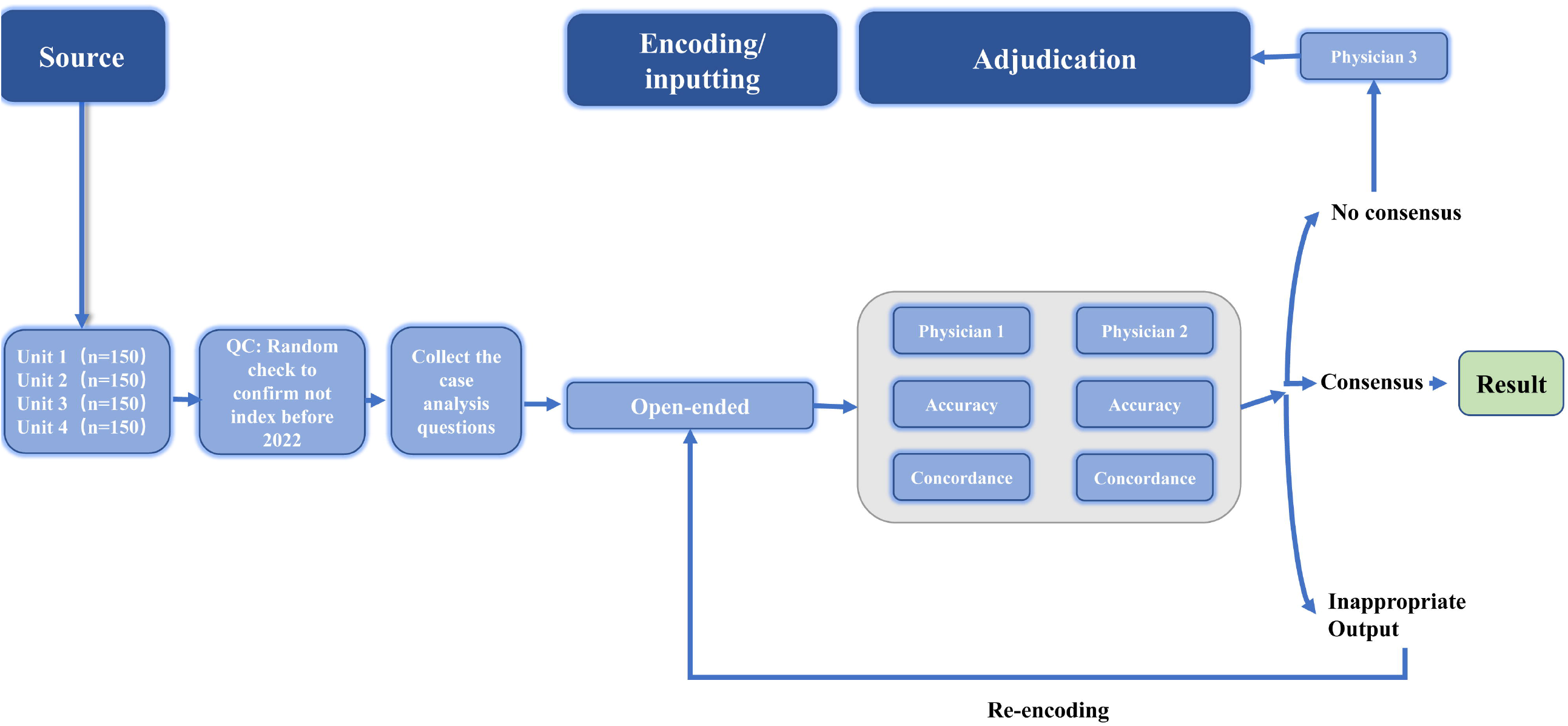
Schematic of workflow for sourcing, encoding, and adjudicating results. The 600 questions were categorized into 4 units. The accuracy of the open-ended encoded questions was evaluated, while the answer with forced justification encoded questions were also assessed for the accuracy, concordance. The adjudication process was carried out by two physicians, and in case of any discrepancies in the domains, a third physician was consulted for adjudication. Additionally, any inappropriate output was identified and required re-encoding.

### Translation

To evaluate if translating questions from Chinese to English could enhance ChatGPT’s performance, we utilized ChatGPT to translate unencoded case analysis questions. We then assessed ChatGPT’s performance on the translated exam by comparing its responses to standard answers and calculating its scores. We compared the scores obtained from the original questions to those from the translated questions and employed the chi-square test to determine performance improvement.

## Result

### ChatGPT passed Chinese NMLE with a high score

In the Chinese NMLE, 442 (73.67%) out of 600 items were correctly answered by ChatGPT, which is significantly higher than the passing threshold (360) defined by official agencies.

The score of each unit is shown in Fig 2. ChatGPT’s performance varied across the four units of questions, with a highest accuracy in Unit 4 (76.0%), followed by Unit 3 (74.7%), Unit 1 (74.0%) and Unit 2 (70.0%), while there was no statistically difference among four units (χ^2^ = 0.66, p = 0.883).

**Fig 2.**
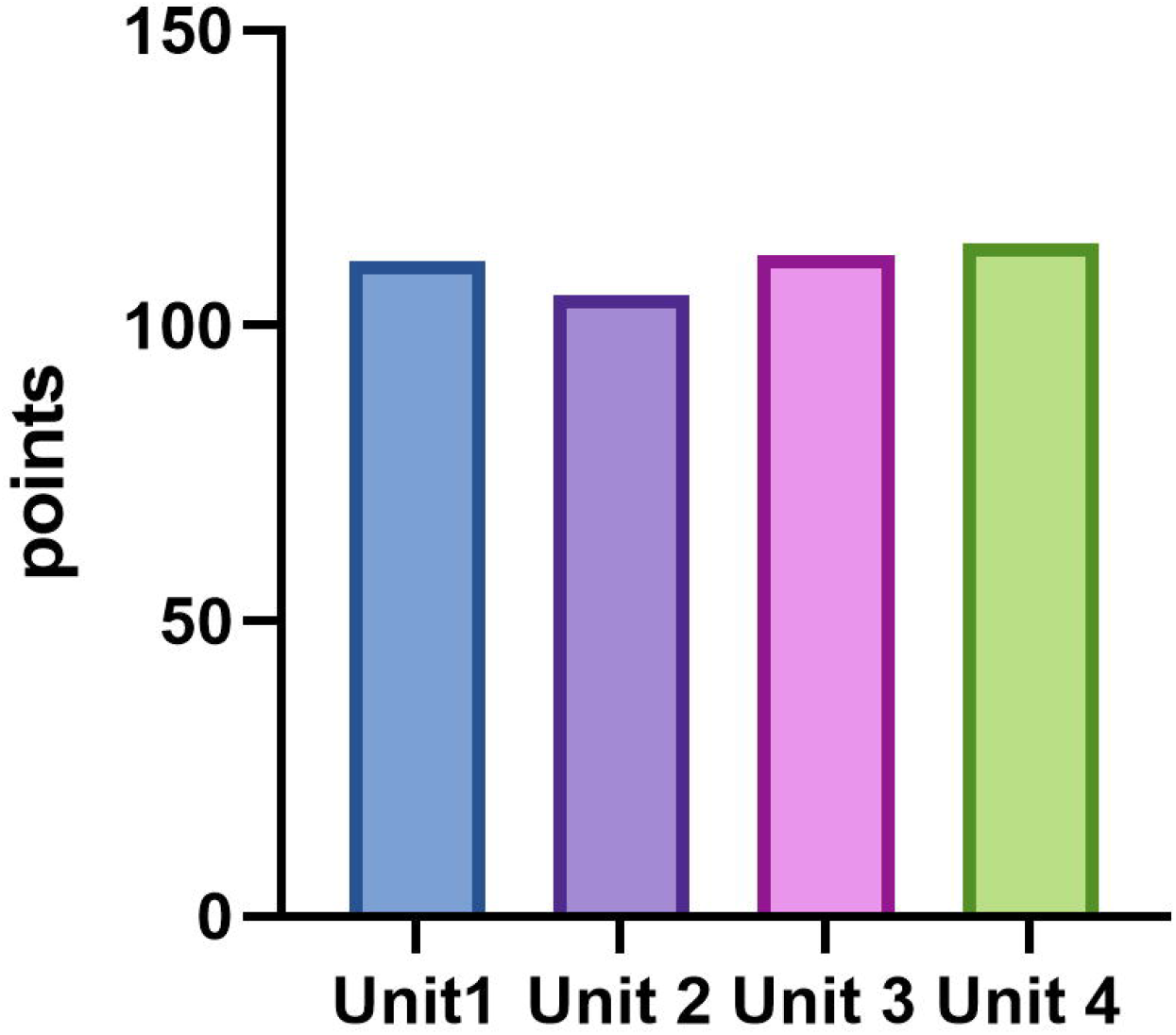
Score of ChatGPT4 on Chinese National Medical Licensing Examination before encoding. ChatGPT’s outputs from Units 1, 2, 3, and 4 were scored for each unit.

### ChatGPT’s performance declines when handling encoded questions

Test questions were encoded as open-ended for case analysis questions, simulating scenarios where a student poses a common medical question without answer choices or a doctor diagnoses a patient based on multimodal clinical data (e.g., symptoms, history, physical examination, laboratory values). The accuracy was 40.5%, 60.3%, 42.3%, and 34.2% for Units 1, 2, 3, and 4, respectively (**Fig 3A**). Compared to the original questions, the accuracy of the encoded questions decreased by 40.5%, 9.7%, 32.4%, and 41.8% for Units 1, 2, 3, and 4, respectively (**Fig 3B**).

**Fig 3.**
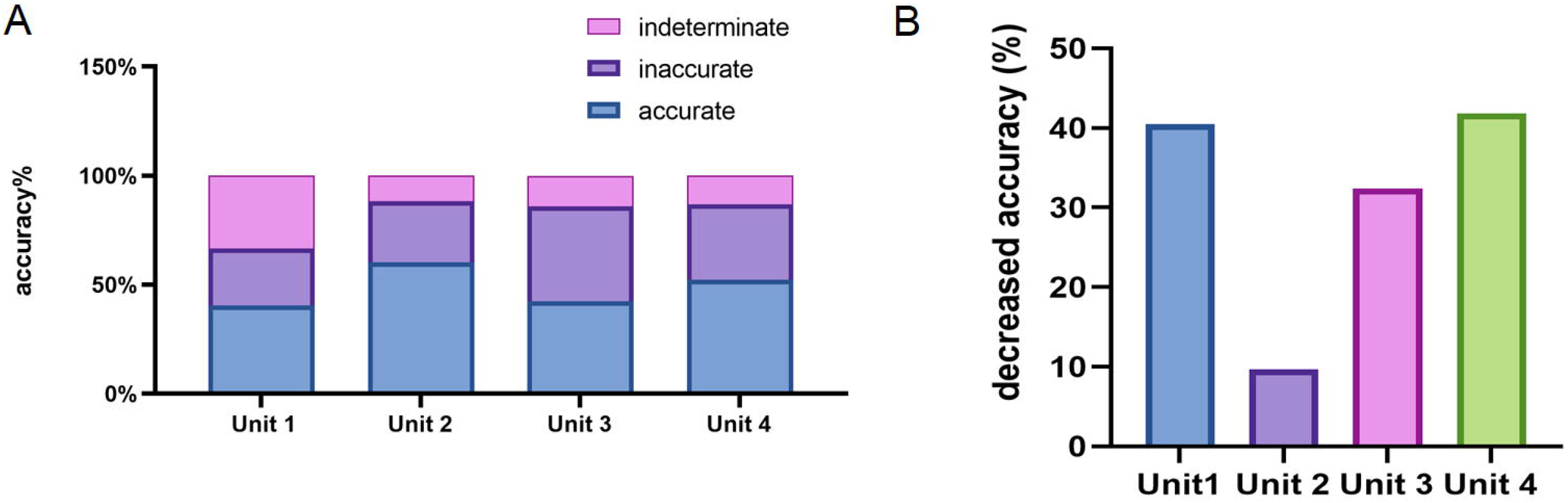
Accurancy of ChatGPT4 on Chinese National Medical Licensing Examination before encoding. ChatGPT’s outputs for Units 1, 2, 3, and 4 were evaluated as accurate, inaccurate, or indeterminate using the scoring system outlined in S2 Data after encoding. (A) Assessment of accuracy for open-ended question encodings. (B) Reduced accuracy of encoded questions across Units 1, 2, 3, and 4.

These findings demonstrate that while ChatGPT’s ability to answer questions in Chinese as applied to common medical situations is commendable, there is still room for improvement. During the adjudication stage, physician agreement was good for open-ended questions (with a κ range from 0.83 to 1.00).

#### ChatGPT demonstrates high internal concordance

Concordance is a measure of the agreement or similarity between the option selected by AI and its subsequent explanation. The results indicated that ChatGPT maintained a >75% concordance across all questions, and this high level of concordance was consistent across all four units (**Fig 4**). Furthermore, we examined the concordance difference between correct and incorrect answers, discovering that concordance was perfect and significantly higher among accurate responses compared with inaccurate ones (85% vs. 59.5%, p<0.005) (**Fig 4**).

**Fig 4.**
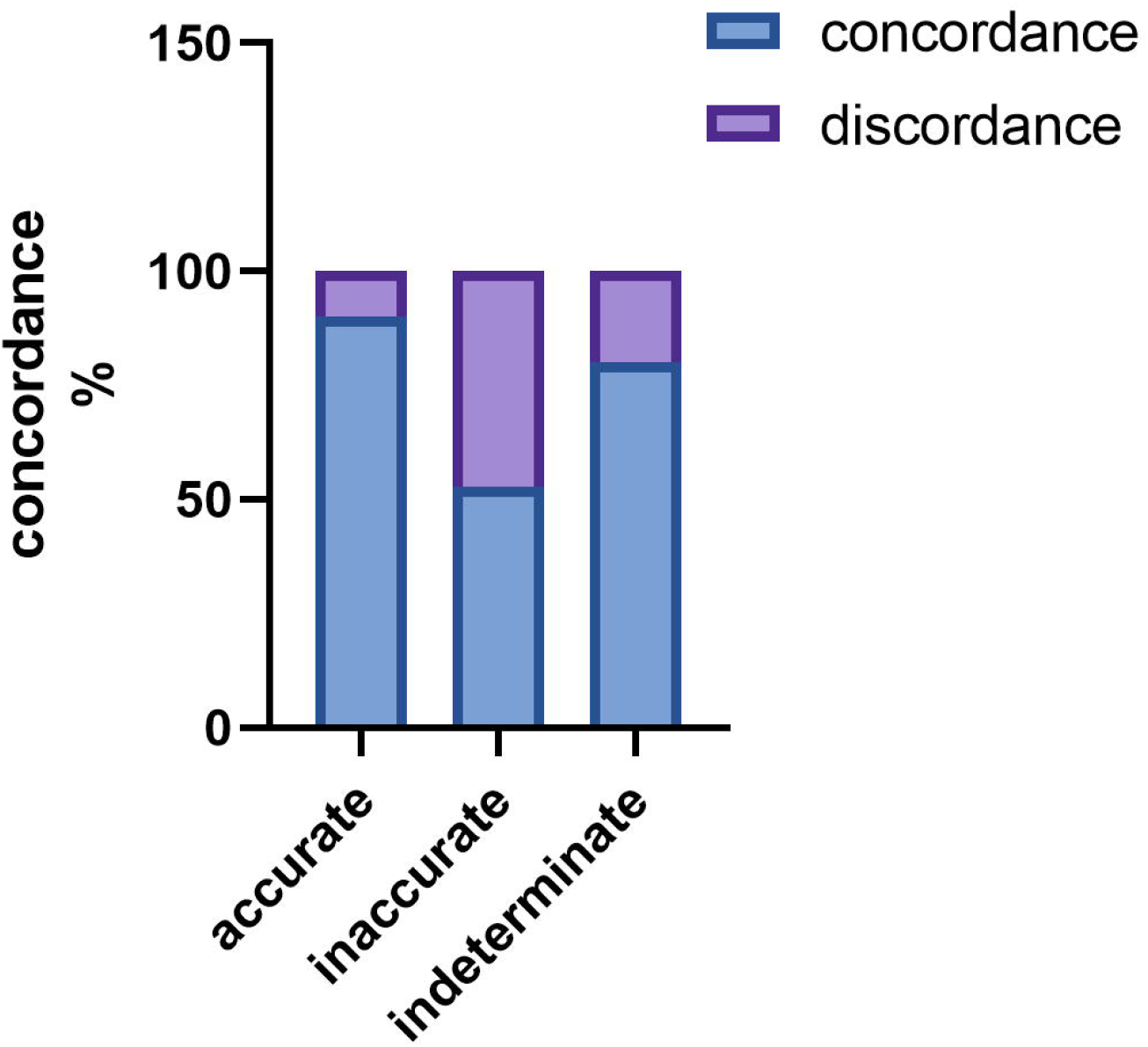
Concordance of ChatGPT4 on Chinese National Medical Licensing Examination after encoding. For Units 1, 2, 3, and 4 after encoding, ChatGPT’s outputs were evaluated as concordant or discordant, based on the scoring system detailed in S2 Data. This figure demonstrates the concordance rates stratified between accurate, inaccurate, and indeterminate outputs, across all case analysis questions.

These findings suggest that ChatGPT exhibits a high level of answer-explanation concordance in Chinese, which can be attributed to the strong internal consistency of its probabilistic language model.

### Translating the input into English may not improve ChatGPT’s performance

After translating the original case analysis questions in Chinese into English to explore a potential way to improve ChatGPT’s performance, the improvement for Units 1, 2, 3, and 4 was minimal, with only one point gained in each unit. The total number of correct answers increased from 256 to 260. The accuracy improvement for translated case analysis questions was subtle (χ^2^ = 0.1206, P =0.728). This suggests that ChatGPT’s performance when facing questions in Chinese may not be improved by translating them into English, and solely building a database in English while translating other languages into English may not be an effective approach.

## Discussion

In present study, we firstly investigated ChatGPT’s performance on the Chinese NMLE. Our findings can be summarized under two major themes: (1) ChatGPT’s score is satisfactory but requires improvement when addressing questions posed in the Chinese language; and (2) Translation into English showed slight performance improvement. This study provides new evidence for the ability of ChatGPT in medical education and clinical decision-making within the Chinese context, offering valuable insights into the applicability of AI language models for non-English medical education settings and laying the groundwork for future research in this area.

### ChatGPT’s performance in the Chinese NMLE is acceptable, yet further improvement

In the Chinese NMLE, ChatGPT achieved a score of 442 (73.67%), exceeding the passing requirement of 360 points for the Chinese language. In the 2022 NLME, the average score of 65 medical students was 412.7 (68.7%), with a minimum score of 295 (49.2%) and a maximum score of 474 (79.0%). According to the statistics, the national pass rate for the exam in 2022 was 55%. When compared to medical students who have undergone a traditional 5-year medical education and a one-year internship, ChatGPT’s performance is currently satisfactory, however, there is still potential for improvement. Several underlying reasons may be responsible. 1) Limitations in training data: If ChatGPT’s training data contains less information about the Chinese medical field, its performance when handling Chinese medical questions could be impacted, resulting in a lower accuracy rate for such queries. 2) Knowledge updates: With a knowledge cutoff date in September 2021, the most recent developments in the Chinese medical field may not have been adequately learned by the model, affecting its accuracy when answering Chinese medical questions.

### ChatGPT’s accuracy can be improved by addressing data limitations, refining its architecture, and using domain-specific knowledge

Moreover, we observed that outputs with high accuracy exhibited high concordance, while lower accuracy was associated with reduced concordance. Consequently, we speculate that ChatGPT’s inaccurate responses primarily arise from missing information, leading to indecision in the AI rather than adherence to an incorrect answer. Language models like ChatGPT are built on vast amounts of text, and their accuracy depends on the quality and diversity of their training data^11^. When the model encounters scenarios with limited or underrepresented data, its performance may suffer, leading to indecision or inaccurate responses. To address this issue, one could consider expanding the training data to cover a broader range of contexts or refining the model’s architecture to handle uncertainty more effectively. Additionally, incorporating domain-specific knowledge and data sources can help improve the model’s performance in specialized areas.

### ChatGPT performs best in English, with accuracy affected by translation issues and data limitations in other languages

ChatGPT’s performance may differ across different language and this variation can be attributed to factors such as the quality and quantity of training data available in different language^12^. Among these languages, a better performance of ChatGPT with English task descriptions is reported^12^. However, we observed a slight improvement in ChatGPT’s performance after translating questions into English, suggesting that relying solely on English databases or translations might not be the an effective approach in improving the abilities for medical tasks. Several potential reasons may be responsible: 1) Translation limitations: When translating questions, some nuances or specific terms may be lost or inaccurately translated, which could impact the AI’s understanding and subsequently its performance^13^. Additionally, some languages may have unique expressions or cultural context that are difficult to convey accurately in English, leading to potential misunderstandings or misinterpretations. 2) AI model’s abilities: Language models like ChatGPT rely on the quality and quantity of training data available in each language. If the model has been trained extensively with English data, it may perform better when handling English text. For other languages, the AI model’s performance could be affected by insufficient or lower-quality training data, leading to less accurate responses.

Regarding the best-performing languages, according to the answer of ChatGPT, English typically yields the highest accuracy since most training data is in English. Other widely spoken languages with a substantial number of online resources, such as Chinese, Spanish, French, and German, may also exhibit relatively better performance in terms of accuracy. However, these results may vary depending on the model and specific task. To assess the AI model’s performance for a particular language, a targeted evaluation might be necessary.

### GPT-4 shows progress, but addressing healthcare standards, ethics, and culture is crucial for AI integration in medicine

ChatGPT-3.5 achieved near-passing threshold accuracy of 60% on the United States Medical Licensing Exam^7^. Furthermore, our previous study also showed a similar performance by ChatGPT-3.5 on the Clinical Medicine Entrance Examination for Chinese Postgraduates (scored 153.5/300, 51%) in Chinese language^14^. In the present study, a significant higher score was found by GPT-4 (scored 442/600,73.6%). This improvement may be attributed to differences in model sizes and training data. GPT-4’s larger model size enables it to handle more complex tasks and generate more accurate responses due to its extensive training dataset, broader knowledge base, and improved contextual understanding^15^. On the other hand, Chinese medical licensing exams have many common-sense questions and fewer case analysis questions than United States Medical Licensing Exam, which may be another reasons for the relatively high pass rates.

Despite the promising potential of AI in medicine, it also faces several challenges. The development of standards for AI use in healthcare is still required^16, 17^, encompassing clinical care, quality, safety, malpractice, and communication guidelines. Moreover, the implementation of AI in healthcare necessitates a shift in medical culture, posing challenges for both medical education and practice. Ethical considerations, such as data privacy, informed consent, and bias prevention, must also be addressed to ensure that AI is employed ethically and for the benefit of patients.

## Limitations

Several limitations should be noted. Firstly, the clinical tasks are highly complicated, the exams cannot fully stimulate the problems in clinical practices. Secondly, the limited input sample size may preclude us performing the depth and range of analyses, which potentially limiting the generalizability of findings. Thus, before large-scale applications of Large Language Model-based AI in medical education or clinical practice, their utility should be further studied in real-world condition.

## Conclusion

ChatGPT demonstrated impressive performance in the Chinese NMLE in Chinese language, exceeding the passing threshold and exhibiting high internal consistency. Nevertheless, its performance waned when faced with open-end encoded questions. Translation into English did not substantially boost its performance. The findings emphasize the ChatGPT’s ability of comprehensible reasoning n medical education and clinical decision-making in Chinese.

## Data Availability

All data produced in the present study are available upon reasonable request to the authors

## Data statement

All data generated or analyzed during this study are included in this published article [and its supplementary information files].

## Acknowledgments

We acknowledge the ChatGPT for polishing our manuscript.

## Author contributions

Xiao Liu was responsible for the entire project and revised the draft. Changchang Fang, Jitao Ling, Jing Zhou, Xiaoling Liu, Yixuan Chen, Zhichen Zhu and Yuan Jiang performed the study selection, data extraction, statistical analysis, and interpretation of the data. Changchang Fang and Xiao Liu drafted the first version of the manuscript. All authors participated in the interpretation of the results and prepared the final version of the manuscript.

## Declarations

Ethics approval This is a systematic review and meta-analysis. No ethical approval is required.

