## supplemental table for "How does ChatGPT4 preform on Non-English National Medical Licensing Examination? An Evaluation in Chinese Language"

**Data Supplement**

**Table S1: The original question**

| Unit 1 | https://drive.google.com/file/d/1aLFag7Z40khalnKckfZR6DYp2TRjWy46/view?usp=share_link |
| --- | --- |
| Unit 2 | https://drive.google.com/file/d/1xElTkdJ8y33WSAmOq0voezaxkTg9OXKP/view?usp=share_link |
| Unit 3 | https://drive.google.com/file/d/1gkbdWjw1imp2OI-AIvs8Ycp1KcH_iiPZ/view?usp=share_link |
| Unit 4 | https://drive.google.com/file/d/1NU1MvFMm4d42zMNpmUlyMk7yyHuWly5x/view?usp=share_link |

**Table S2：Adjudication criteria for accuracy, concordance.**

| Accurate:1. Provide the answer accurately.  2.When the judge determines that there is not a unique answer, the AI outputs multiple choices, among which contains the correct answer, and the other options are also completely correct. | Concordant: Explaining affirms the answer |
| --- | --- |
| Inaccurate 1. No answer is provided  2.An incorrect answer is provided  3.Multiple answers are provided, among which there is an incorrect answer, even if the correct answer is included. | Discordant: Explanations are contradictory. |
| Indeterminate: 1.AI output is not a single answer election.  2.When the judge determines that there is a unique answer, AI output provides multiple choices.  3.AI believes that there is not enough information." |  |

**Table S3：Kappa statistic for interrater agreement between adjudicating physicians.**

|  | Accuracy | | Concordance | |
| --- | --- | --- | --- | --- |
|  | Cohen κ | n | Cohen κ | n |
| Unit 1 | 1 | 42 | 1 | 0.88 |
| Unit 2 | 0.88 | 68 | 1 | 68 |
| Unit 3 | 0.83 | 71 | 0.97 | 71 |
| Unit 4 | 0.85 | 79 | 1 | 79 |

**Table S4：Crosstab for evaluating translation effectiveness**

|  | |  | | Sum |
| --- | --- | --- | --- | --- |
|  |  | Accurate | Inaccurate |  |
|  | Original questions | 197 | 63 | 260 |
|  | Translated questions | 201 | 59 | 260 |
| Sum | | 398 | 122 | 520 |
